# The impact of non-invasive prehabilitation before surgery on emotional well-being in neuro-oncology patients: Insights from the Prehabilita project

**DOI:** 10.64898/2026.04.08.26350382

**Authors:** Nuria Brault-Boixader, Alba Roca-Ventura, Selma Delgado-Gallén, Edgar Buloz-Osorio, Ruben Perellón-Alfonso, Chi Hung Au, David Bartrés-Faz, Álvaro Pascual-Leone, José Maria Tormos Muñoz, Kilian Abellaneda-Pérez, Prehabilita Working Group

**Affiliations:** Institut Guttmann, Institut Universitari de Neurorehabilitació adscrit a la UAB, Badalona, Spain; Universitat Autònoma de Barcelona, Bellaterra, Spain; Fundació Institut d’Investigació en Ciències de la Salut Germans Trias i Pujol, Badalona, Barcelona, Spain; Departament de Medicina, Facultat de Medicina i Ciències de la Salut, i Institut de Neurociències, Universitat de Barcelona, Barcelona, Spain; Institut d’Investigacions Biomèdiques August Pi I Sunyer (IDIBAPS), Barcelona, Spain; Unitat de Neurociència Cognitiva, Institut de Neurociències, Universitat de Barcelona, Barcelona, Spain; NeuroLinks Clinic: 6010 Brickyard Rd #202, Nanaimo, British Columbia, Canada; Department of Psychiatry, University of British Columbia, Vancouver, British Columbia, Canada; Wolk Center for Memory Health & Marcus Center for Aging Research, Hebrew SeniorLife, Boston, MA, USA; Department of Neurology, Harvard Medical School, Boston, MA, USA; Centro de Investigación Traslacional San Alberto Magno, Universidad Católica de Valencia San Vicente Mártir, Valencia, Spain; Laboratori de Neurociència Cognitiva, Departament de Psicologia, Universitat de Lleida, Lleida, Spain; Institut de Recerca Biomèdica de Lleida – Fundació Dr. Pifarré, IRBLleida, Lleida, Spain

**Keywords:** psycho-oncology, emotional well-being, quality of life, emotional distress, neuro-oncology, prehabilitation

## Abstract

Prehabilitation (PRH) is a preoperative process aimed at optimizing patients’ functional capacity to improve surgical outcomes and overall well-being. While its physical and cognitive benefits are increasingly documented, its emotional impact, particularly in neuro-oncology patients, remains less explored. This study assessed the psychological effects of a PRH program on 29 brain tumor patients. The primary outcome, emotional well-being, was measured using quality of life and emotional distress metrices. Secondary outcomes included perceived stress levels and control attitudes. Additionally, qualitative data from structured interviews provided further insights into the psychological effects of the intervention. The results indicated significant improvements in quality of life and reductions in emotional distress, particularly among women. While perceived stress levels remained stable, control attitudes showed an increase. Qualitative analysis further highlighted the positive changes in the control sense and identified additional factors, such as the importance of social support sources during the PRH process. Overall, these findings suggest that PRH interventions play a significant role in enhancing emotional well-being among neuro-oncological patients in the preoperative phase. These results underscore the importance of implementing comprehensive and personalized PRH approaches to optimize clinical status both before and after surgery, thereby promoting sustained psychological benefits in this population. This study is based on data collected at Institut Guttmann in Barcelona in the context of the Prehabilita project (ClinicalTrials.gov identifier: NCT05844605; registration date: 06/05/2023).

## 1. Introduction

Brain and central nervous system (CNS) cancers represent a complex group of neoplasms with high morbidity, disability, and mortality (1). According to Global Cancer Statistics 2022, there were 321,476 new cases of brain and CNS cancers worldwide, leading to 248,305 deaths that year (2). In Spain, where the present investigation was conducted, an estimated 2,331 cases in men and 2,080 in women were diagnosed in 2023 (3). Brain cancer treatment habitually involves a multifaceted approach that may encompass surgery, chemotherapy, and radiation therapy (4). Surgical resection constitutes a central treatment strategy, requiring a delicate balance between maximizing tumor removal while preserving critical neurological function. Patients’ survival and quality of life (QoL) are associated with both the extent of tumor resection and postoperative neurological deficits (4–6), respectively.

From an emotional well-being perspective (EWB), being diagnosed with a brain tumor can be particularly distressing as patients face the dual challenge of life-threatening disease and the risk of profound neurological impairment (7). This diagnosis is frequently accompanied by emotional strain, cognitive dysfunction, fatigue, fear, memory or concentration difficulties, worry and a sense of uncertainty that extends beyond the patient to their families (7–9).

Clinically significant distress is experienced by almost half of cancer patients as a consequence of their diagnosis and subsequent treatment (10). Brain tumor patients, specifically, tend to experience higher distress levels compared to those diagnosed with non-CNS tumors (11,12).

The perioperative period is especially critical, as the combination of preoperative stress, potential postoperative complications, and prolonged recovery times can leave patients feeling vulnerable and overwhelmed (13). Many also suffer from the anticipatory grief – a form of mourning for losses that have not yet occurred (14).

In recent years, prehabilitation (PRH) interventions have emerged as strategies to optimize patients’ functional and neurological status by promoting the reorganization of brain networks involved in peritumoral areas, potentially improving postoperative recovery and outcomes (15). Although PRH initiatives have shown promise in improving patients’ physical condition and readiness for surgery, they often lack a strong focus on EWB. From this perspective, previous studies (largely drawn from non-neuro-oncology populations) suggest that PRH may help redirect patients’ attention away from cancer-related worries by providing a constructive and purposeful focus. These programs can also effectively foster a sense of active participation and control over the patient’s own healthcare, which can be empowering during treatment (15,16). Patients who report a stronger sense of perceived control, or an internal locus of control, tend to experience lower levels of depression and anxiety, along with better QoL at the time of their cancer diagnosis (17,18). Reflecting this, there is growing interest in interventions specifically designed to enhance psychological resilience of patients prior to surgery (10). In this context, the work of Powell and colleagues (15), provides a valuable framework to investigate the psychological effects of PRH in non-neuro-oncology patients.

The present investigation aims to specifically examine the impact of the Prehabilita project (19), a PRH program conducted at Institut Guttmann in Barcelona, on the EWB of its participants. The Prehabilita project is a research initiative aimed at optimizing tumor resection while preserving functional outcomes. It focuses on promoting brain functional reorganization away from the peritumoral area. This is achieved by leveraging the brain’s plasticity through non-invasive neuromodulation techniques, combined with intensive cognitive and motor training programs. However, the psychological impact of our program has thus far not being systematically investigated. Consequently, the main goal of the present study was to evaluate the impact of our PRH program on EWB, considering as the main outcomes QoL and emotional distress. Our secondary goal was to identify potential psychological mechanisms influencing EWB changes during PRH.

## 2. Materials and methods

### 2.1 Participants

This study is based on data collected at Institut Guttmann in Barcelona in the context of the Prehabilita project (ClinicalTrials.gov identifier: NCT05844605; registration date: 06/05/2023) and includes 29 participants (11 women, 18 men; mean age = 56.27, SD = ±11.58; range = 30-77; see Table 1), who completed the PRH program as well as QoL and emotional distress questionnaires. Of note, participants were classified by biological sex (female or male) based on medical records. This classification aligned with the gender the participants identified with (women and men, respectively). Therefore, gender identity was used in this study. Inclusion and exclusion criteria for this study are described in greater detail elsewhere, as well as safety and feasibility (19–21), but briefly: Adults were referred to by neurosurgery units across Catalonia. Those requiring neurosurgery for brain tumors in eloquent areas for motor and language function, were deemed eligible for PRH at Institut Guttmann in Barcelona: Neurorehabilitation Hospital. Exclusion criteria included contraindications to transcranial magnetic stimulation (TMS) or magnetic resonance imaging (MRI, i.e. metal implants or claustrophobia), unstable medical conditions (such as heart disease, asthma or respiratory conditions, high blood pressure, osteoporosis, or diabetes), substance abuse history, and severe musculoskeletal or cognitive disorders impacting the intervention. Moreover, participants needed to comprehend the study’s purpose, provide written informed consent, and agree on attending a minimum of 10 PRH sessions (with a maximum of 20). The intervention comprised a daily regimen of neuromodulation, via non-invasive brain stimulation (NIBS), immediately followed by intensive functional training (motor and/or cognitive). Neurosurgery was performed soon after completing the 10-20 sessions, to make sure that the neuroplastic changes and functional adaptations induced persist throughout the surgery. Please refer to Figure 1 for an overview of the process timeline. A more detailed description of the PRH procedures can be found in the protocol paper by (20). Thirteen subjects were taking medication during the Prehabilita project, specifically psychotropic medications, including anxiolytics (benzodiazepines), antidepressants (SSRIs), and other types of antidepressants (See Table 1). However, none of them changed their medication during the course of the project.

**Figure 1.**
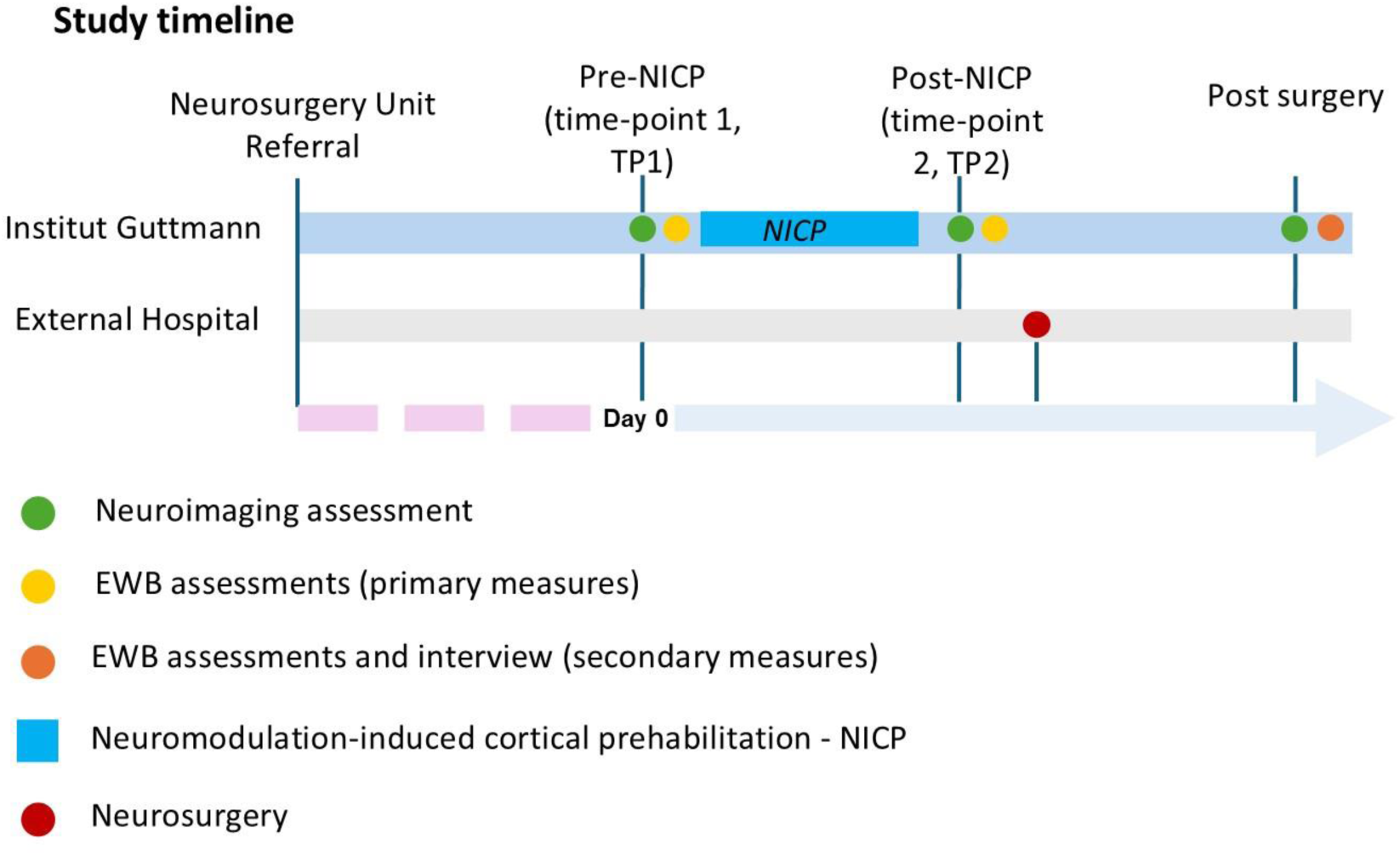
Study timeline, where emotional well-being assessments are depicted in yellow for primary measures (EORTC-QLQ, EORTC QLQ-BN20 and HADS) and in orange for secondary measures (PSS-4, CAS and three-question interview).

**Table 1.** Participant main demographic and clinical variables. Percentages are based on the total sample size (N = 29). Note: Psychiatric medication includes antidepressants and anxiolytics.

| **Variable** | **Category or Measure** | **Value** |
| --- | --- | --- |
| **Age (years)** |  |  |
| | Mean (SD) | 56.27 (±11.58) |
|  | Range | 30–77 |
| **Gender** |  |  |
|  | Women | 11 (37.9%) |
|  | Men | 18 (62.1%) |
| **Brain tumor type** |  | **N° of participants** |
|  | Glioblastoma | 13 (44.82%) |
|  | Astrocytoma | 5 (17.24%) |
|  | Oligodendroglioma | 3 (10.34%) |
|  | Meningioma | 3 (10.34%) |
|  | Cavernoma | 2 (6.89%) |
|  | Ganglioglioma | 1 (3.44%) |
|  | Gliosarcoma | 1(3.44%) |
|  | Carcinoma | 1(3.44%) |
| **Psychopharmacologic treatment** |  |  |
|  | Psychiatric medication | 13(44.82%) |

### 2.2 EWB assessments

#### 2.2.1 EWB primary measures

In this study, we analyzed data from the first two time points (TP1 and TP2) of the whole PRH protocol (i.e., before and after neuromodulation-induced cortical prehabilitation - NICP). The questionnaires used in this study to explore the main EWB outcomes of the intervention were the European Organization for Research and Treatment of Cancer Quality of Life Questionnaire (EORTC - QLQ) and the Hospital Anxiety Depression Scale (HADS). Both questionnaires were administered by a trained neuropsychologist.

The EORTC-QLQ is a standardized tool developed to assess the health-related QoL of cancer patients participating in clinical trials. The instrument consists of a core questionnaire (EORTC QLQ-C30), which evaluates general QoL aspects in cancer patients, as well as disease-specific and symptom-specific modules tailored for cancer types and treatment modalities (22). The core questionnaire contains 30 items and assesses: global health status and QoL; functional domains: physical, role, cognitive, emotional, and social functioning and financial difficulties; symptom domains: fatigue, pain, nausea/vomiting, dyspnea, insomnia, appetite loss, constipation, diarrhea.

Furthermore, the EORTC QLQ-C30 includes supplementary modules designed for specific cancer types. As a complement to this, we also used the EORTC QLQ-BN20, a brain tumor-specific module of the EORTC QLQ-C30, designed to assess the impact of the tumor and its treatment on symptoms, functions, and health-related QoL (i.e., patient’s overall well-being, emotional state, and social interactions), in both clinical trials and clinical practice. This 20-item module evaluates key domains such as future uncertainty, visual disorder, motor dysfunction, and communication deficit. In this questionnaire, higher scores indicate poorer functioning (22,23).

The HADS (24) questionnaire was used to capture emotional distress in its two main components: anxiety and depression. Therefore, it consists of two subscales: HADS-A for anxiety, and HADS-D for depression. Each subscale comprises seven items with a four-point Likert scale response format. Scores range from 0 to 21 in each subscale, with higher scores indicating more anxiety or depression. The HADS general score cutoff (A+D) for meeting clinical criteria is ≥15. This cut-off point is consistent with the original recommendations of (24) and have been validated in various populations, including oncology patients in Spain.

#### 2.2.2 EWB secondary measures

After completion of the PRH program, we conducted a phone-based enrichment evaluation to gather further information about patients’ affective and cognitive experiences during the program.

This evaluation included two validated questionnaires to assess locus of control and perceived stress, namely the 4-item Control Attitudes Scale (CAS) and the 4-item Perceived Stress Scale (PSS-4), respectively, as well as a brief structured interview to collect additional qualitative data. Both questionnaires were administered by a trained psychologist. Although this enrichment evaluation took place only after the program, participants were explicitly asked to reflect on their EWB both before and after the intervention. Thus, the data capture included a retrospective assessment of the pre-intervention state. A total of 14 participants, who were able to respond and provided informed consent, completed these additional measures.

The 4-item CAS (25,26), consists of 4 items that measure an individual’s perception of control over their health or illness. The CAS is especially relevant in examining how control attitudes influence psychological outcomes such as emotional distress, coping strategies, and adherence to medical treatments. Respondents rate these items on a Likert scale (e.g., 1 = very little control, 7 = very high control). In this instrument, higher scores indicate a greater sense of perceived control.

The PSS-4 (27) is an ultra-brief self-report measure to assess perceived psychological stress. The four items are rated on a 5-point Likert scale (0 = never, 4 = very often). Scores range from 0 to 16, with higher scores indicating higher levels of perceived stress. It has been validated in the United Kingdom, France, and Spain (28).

In addition, we conducted a brief and structured three-question interview to explore participants’ opinions and experiences within our PRH program. The core concept of this interview was to assess how the program influenced participants’ perception of control and the support they received during the process, as well as how these psychosocial variables impacted their EWB. This qualitative questionnaire was adapted from Powell and colleagues (15). The full interview guide is provided in Supplementary File 1.

### 2.3 Data analysis

#### 2.3.1 Quantitative analyses

##### 2.3.1.1 Primary measures: EWB outcomes

QoL was assessed by computing total scores for the EORTC-QLQ and the EORTC-BN20 separately, and emotional distress was assessed using the total score of the HADS scale. For each of these measures, we first conducted repeated-measures ANOVAs, with time (TP1: pre-prehabilitation vs. TP2: post-prehabilitation) as a within-subjects factor, and sex (woman vs. man) as between-subjects factors. Post-hoc comparisons were conducted using Student’s paired-samples t-tests for significant main or interaction effects identified by the repeated-measures ANOVAs.

##### 2.3.1.2 Secondary measures: EWB-related mechanisms

Perceived level of control was calculated by computing the sum of scores of all the CAS scale items and perceived psychological stress was calculated by computing the sum of scores of all the PSS-4 scale items. Due to the small sample size (N = 14), these secondary measures were analyzed without subgroup stratification, as subgroup analyses would lack sufficient statistical power. Additionally, the limited sample size restricted the appropriateness of repeated-measures ANOVA due to concerns about robustness and assumptions regarding normality and variance homogeneity. Therefore, differences in CAS and PSS-4 scores between time points were tested using Student’s paired-samples t-tests. We also explored correlations between CAS, HADS, and EORTC scores using Pearson’s product-moment correlations.

For both primary and secondary measures analyses, when parametric assumptions were not met, we employed nonparametric alternatives: Wilcoxon signed-rank tests for paired comparisons, Welch’s tests for comparing group means, and Spearman’s rank correlation coefficients for associations.

#### 2.3.2 Qualitative analyses

Participant responses to the three questions were transcribed verbatim during the interview. Then, we conducted a qualitative thematic analysis (15,29), to explore patient’s perspectives on the PRH program and its impact on EWB. An inductive coding approach was applied, with line-by-line analysis to identify initial codes (labels or short phrases assigned to segments of qualitative data that represent key concepts, patterns, or themes). The first author (NBB) carried out all coding and thematic development. To enhance trustworthiness, a detailed audit trail was maintained throughout the analytic process, and findings were regularly discussed with the research team to promote reflexivity and reduce potential bias (The audit trail is provided in Supplementary File 2). AI-assisted tools (30) were used to support the organization and clustering of codes with shared meanings into broader themes, which were iteratively refined as new insights emerged.

#### 2.3.3 Control analysis

To further elucidate individual variability in intervention response, we examined whether the spatial proximity of each participant’s TMS target to canonical left dorsolateral prefrontal cortex (DLPFC) coordinates, which are commonly employed in neuromodulation protocols for major depressive disorder, was associated with emotional outcomes (31). Specifically, we investigated whether anatomical proximity to three well-established DLPFC targets (32–34) predicted changes in EWB within our cohort.

To conduct these analyses,___we first calculated the Euclidean distance between each participant’s TMS stimulation coordinate and the three group-defined reference points selected within the DLPFC (see (35)). We then assessed whether this distance was associated with changes in EWB scores using Pearson’s product-moment correlations. Detailed correlation values for each of the three selected DLPFC reference coordinates are provided in Supplementary File 3 and Supplementary Table S1.

## 3. Results

### 3.1 Primary measures: QoL

The analysis of the EORTC-QLQ and EORTC-BN20 data revealed no significant interaction between the within-subject factor “time” (TP1, TP2) and the between-subject factor “sex” on QoL. No baseline differences between groups were observed, *W* (11.07) = -1.71, *p* > .05.

Regarding changes over time, no significant difference was found in EORTC-QLQ scores between TP1 and TP2, *Z* = -0.56, *p* > .05. However, a significant reduction in EORTC-BN20 scores was observed, *Z* = 2.06, *p* = .041, with a medium effect size (*d* = 0.5). On average, scores decreased from TP1 (*M* = 19.63, *SD* = 15.03) to TP2 (*M* = 14.68, *SD* = 11.39). Concretely, 15 out of 25 participants (60%) showed improvement in EORTC-BN20 scores over time (see Figure 2A).

**Figure 2.**
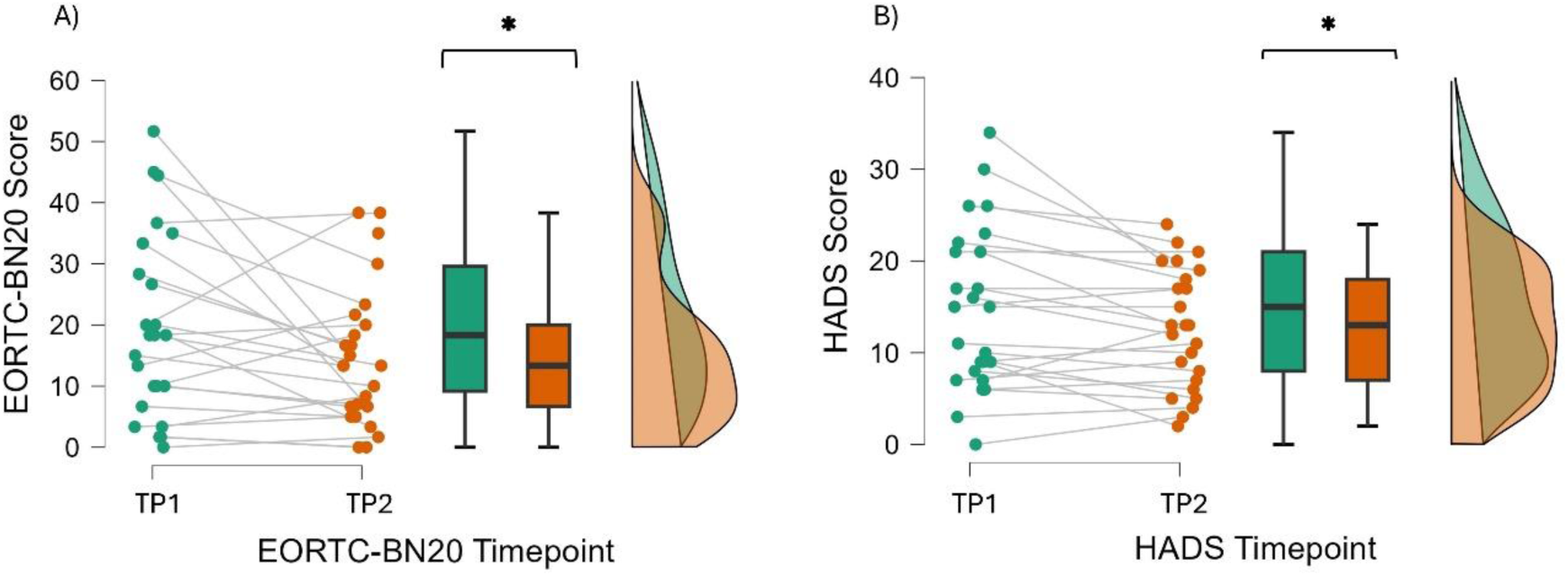
Comparison of the EORTC-BN20 (A) and HADS (B) general score, between TP1 and TP2 timepoints (mean = 14.72 and 12.56, respectively). In the left side of each image, plots are individualized, and in the right side, grouped. Asterisks (*) indicate statistical significance.

### 3.2 Primary measures: Emotional distress

A repeated-measures ANOVA was conducted with “time” (TP1 vs. TP2) as a within-subject factor and “sex” (W vs. M) as a between-subject factor. The analysis revealed a significant main effect of “time”, indicating an overall reduction in HADS scores from TP1 (*M* = 14.72, *SD* = 8.84) to TP2 (*M* = 12.56, *SD* = 6.54), *F*(1, 23) = 22.91, *p* = < .001, η²ₚ = 0.036 (see Figure 2, B). Specifically, 15 out of 25 participants (60%) experienced emotional improvement.

Additionally, the analysis revealed a significant interaction between “time” and “sex”, F(1, 23) = 25.79, *p* < .001, η²ₚ = .04. Post-hoc paired samples t-tests indicated that women exhibited a significant reduction in distress from TP1 to TP2, t(8) = 4.95, p < .001, whereas men showed no statistically significant change, t(15) = -0.29, p = .77 (see Figure 3).

**Figure 3.**
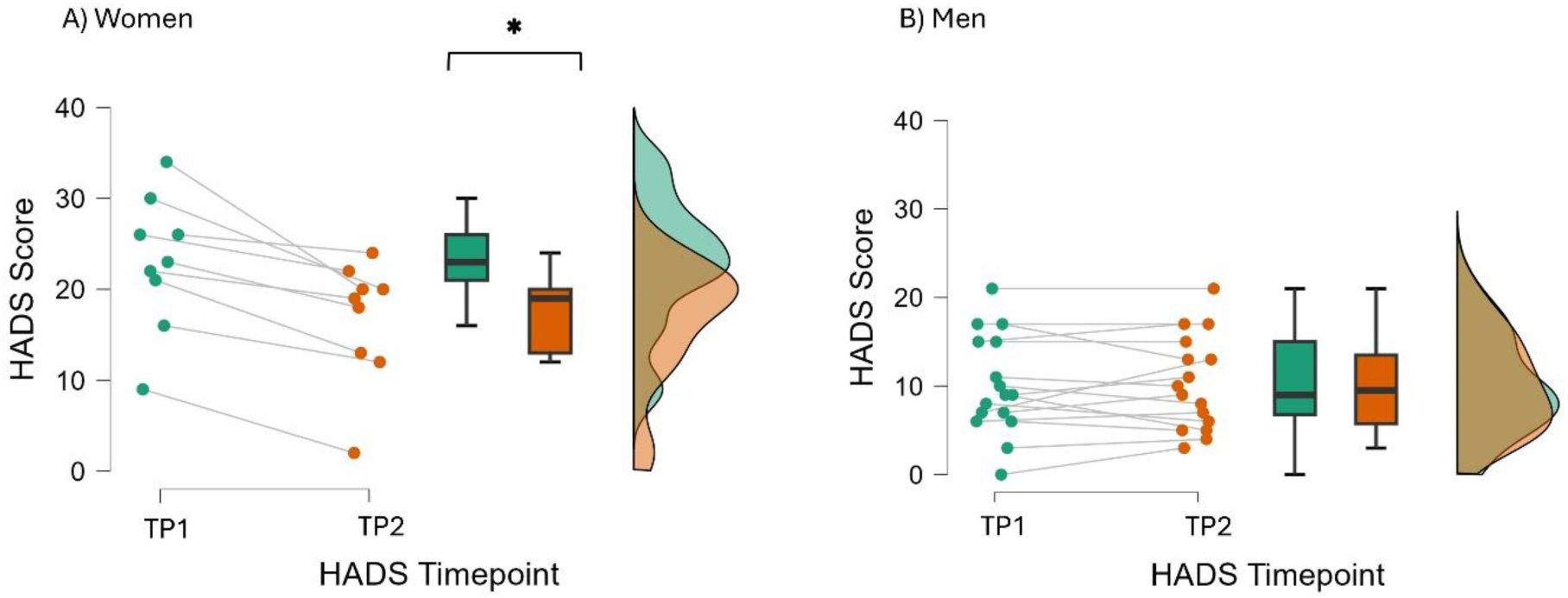
Graphical description of distress change over time in women (A) and men (B). In the left side of each image, plots are individualized, and in the right side, grouped. Asterisks (*) indicate statistical significance.

Finally, from a clinical perspective, 13 of the 25 participants scored above the clinical threshold of 15 on the HADS. Notably, following the intervention, the number of participants above the threshold decreased to 10.

### 3.3 Secondary measures: Locus of control

Analysis revealed a significant difference between CAS TP1 and TP2 timepoints, indicating an increase in patients’ perception of sense of control after the intervention as compared to baseline, *t*(13) = -4.80, *p* <.001 (See Figure 4, A). More specifically, in 13 out of 14 subjects this perception increased, representing 92.85% of the sample.

**Figure 4.**
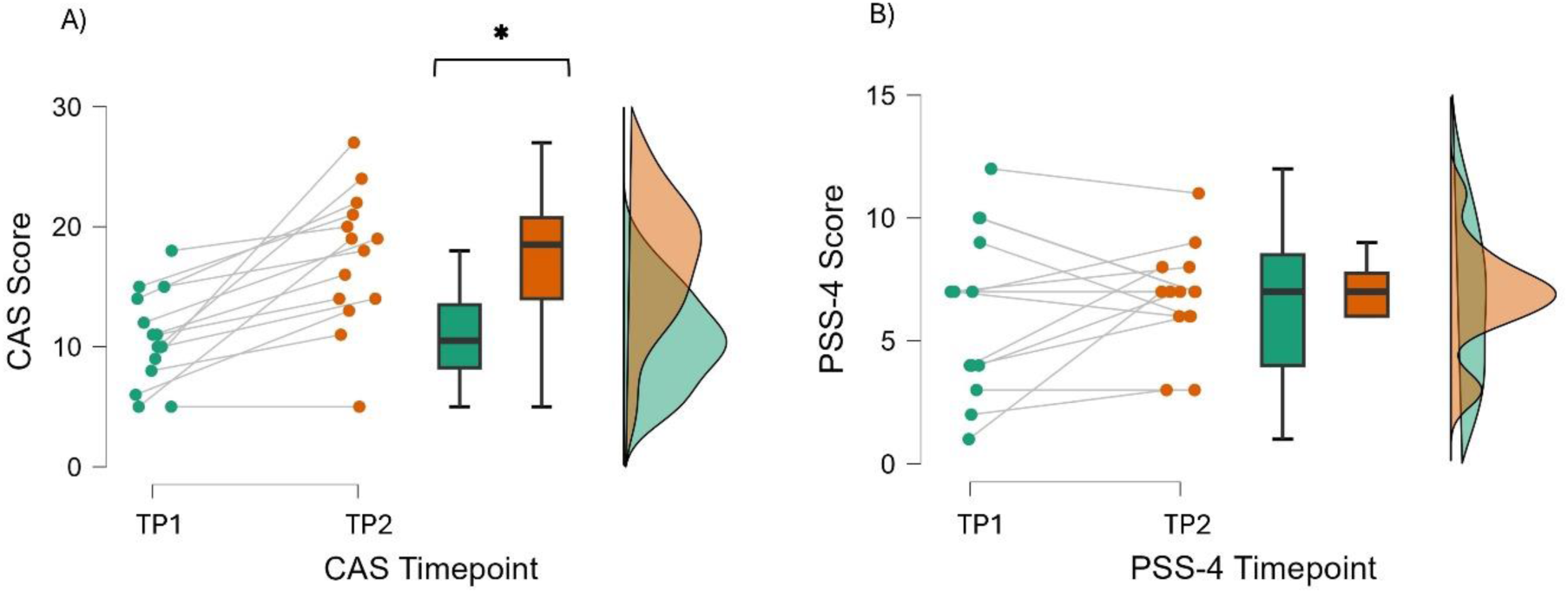
Diagram describing the temporal change of locus of control (A) and perceived stress (B). In the left side of each image, plots are individualized, and in the right side, grouped. Asterisks (*) indicate statistical significance.

When correlating CAS data with EORTC-BN20 and HADS, no significant results were found (rho = 0.136, p=.663; rho = 0.196, p=.707, respectively).

### 3.4 Secondary measures: Perceived stress

No significant differences were observed in general perceived stress when comparing the two timepoints on the PSS-4, *t*(13) = -0.79, *p* = 0.444 (See Figure 4, B).

### 3.5 Qualitative results

Two analytical themes were identified relating to the EWB impact of the PRH program, using data from patient responses: sources of support during PRH and control over medical situation (see Table 2).

**Table 2.** Illustrative quotes from participant interviews, divided by analytical themes: Sources of emotional support during PRH and control over the medical situation.

| **Analytical themes** | **Case n<sup>o</sup>** | **Example** |
| --- | --- | --- |
| Sources of emotional support during PRH | *Case I* | *“I always felt supported, both by the healthcare team and my family”.* |
|  | *Case II* | *“I felt good, loved. Now I feel very good, and I will always be grateful to these women [Prehabilita team], and my friends”.* |
|  | *Case XIII* | *“My hospital, you [Prehabilita team], and my family have all helped me a lot”.* |
| Control over medical situation | *Case V* | *“They also saw my willingness to engage. I always asked a lot of questions, to be aware of what could happen to me and to prepare for possible situations—to anticipate as many scenarios as possible. No matter what happened, I wanted people to know how to act”.* |
|  | *Case VI* | *“Yes, I think so (referring to having more control), because I left there very pleased, with a lot of help and collaboration. I was there for a month before the surgery, it was every day, but the experience was very good”.* |
|  | *Case IX* | *“Prehabilitation reduced the feeling that I had no control, and I was able to face the diagnosis better”.* |

#### Sources of emotional support during PRH

Participants frequently described receiving emotional support from various sources, including family members, friends, colleagues, neighbors, social circles, and patient networks. They also highlighted the strong guidance provided by medical professionals, such as medical doctors, nurse practitioners, psychologists, and the PRH team (see Figure 5). Terms like “supported,” “accompanied,” “lucky,” “safe,” “serenity,” and “comfort” frequently emerged.

**Figure 5.**
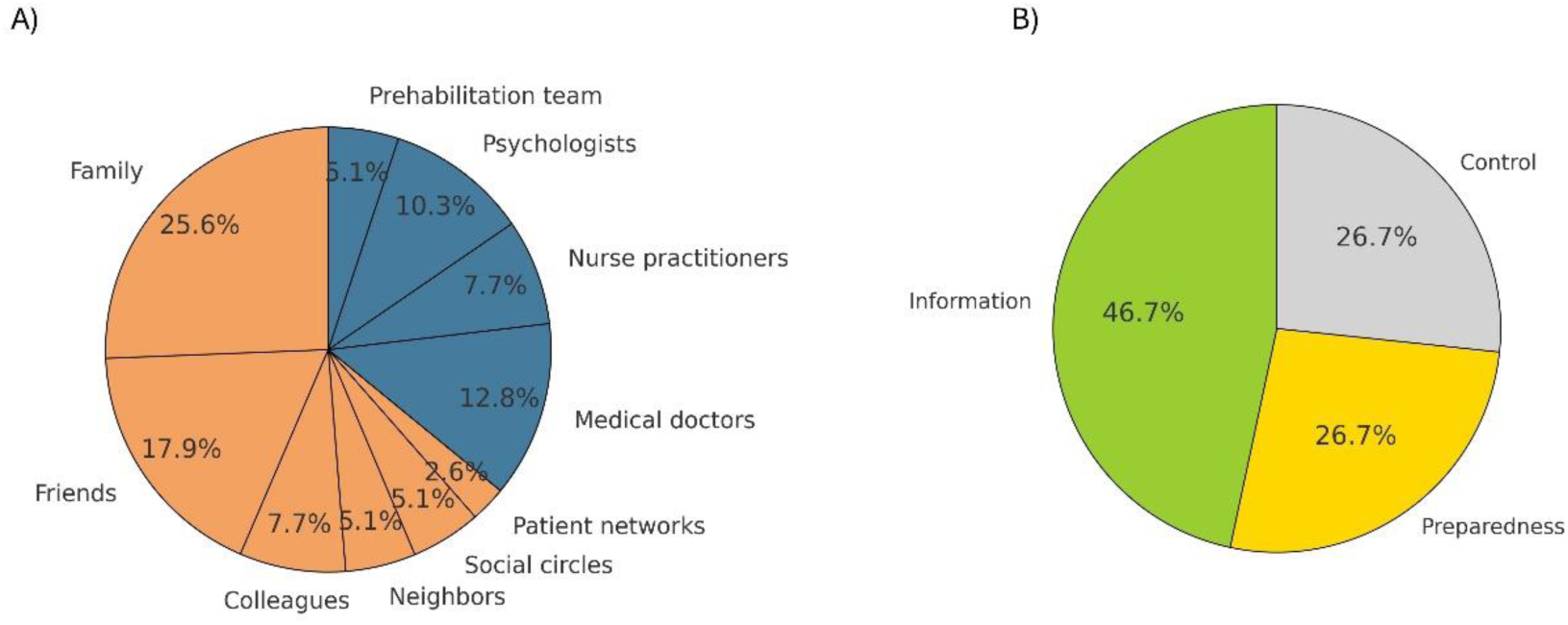
Pie charts showing (A) the most frequently mentioned sources of emotional support during PRH and (B) key aspects contributing to perceived control over the medical situation during PRH. Note: The proportions shown in the pie charts are based on a qualitative frequency analysis of participant narratives. Each category reflects the number of times a concept or theme was explicitly mentioned or clearly implied across the responses. The counts are not literal word frequencies, but rather thematic groupings derived from content analysis to capture the most salient aspects of participants’ experiences.

#### Control over medical situation

Many respondents acknowledge feeling more informed and prepared for surgery due to PRH (see Figure 5). Some responses express concerns about a lack of clear, structured medical information before entering the PRH program. Terms like “serenity”, “information”, “prevention”, and “control” frequently emerged.

## 4. Discussion

The present study evaluated whether a non-invasive PRH program, primarily focused on reducing physical and cognitive sequelae, could also improve QoL and reduce emotional distress in brain tumor patients scheduled for surgery. We observed significant improvements in both QoL and emotional distress following the PRH program. Moreover, women showed greater reductions in emotional distress levels compared to men. Further, the secondary measure of locus control improved post-intervention.

Importantly, although it could be assumed that the increased number of medical appointments and tests before surgery might heighten emotional distress, our findings did not support this concern. This is particularly relevant in relation to anticipatory grief—a type of psychological distress that arises when patients and families begin to mourn expected losses. The perioperative period, often filled with uncertainty and fear, can intensify these emotions (13,14). Our results suggest that the structured support offered through PRH may instead help mitigate this impact by fostering emotional preparedness and a greater sense of control.

Qualitative findings further reinforced the perceived value of the program. Participants commonly reported that PRH enhanced their sense of preparedness and control prior to surgery. Emotional support from family, medical professionals, and community networks emerged as key factors facilitating coping. These results are aligned with previous PRH research conducted in non-CNS cancers, such as Powell et al. (15), who examined PRH and recovery interventions before cancer surgery, finding that patients reported reduced anxiety and increased confidence in coping with treatment.

### 4.1 Impact of PRH on QoL and emotional distress

Existing literature highlights the significant psychological burden of a brain tumor diagnosis due to its potential impact on survival, neurological function, and overall QoL (7,8). Even in the absence of major complications, there is a 20–40% reduction in postoperative physical function and a deterioration in QoL after major surgery in older adults (36). Recent studies reinforce the broader benefits of PRH on postoperative recovery and overall QoL. For instance, Chou et al. (37), noted that cancer PRH programs, both physical and psychological, can improve QoL by enhancing preoperative conditions and promoting better recovery after surgery. Furthermore, McIsaac et al. (38), found that exercise and nutrition-based PRH significantly reduced complications and improved QoL. Moreover, the inclusion of psychosocial support in these interventions further improved recovery outcomes. In line with these findings, our study participants experienced notable and clinically relevant improvement in QoL, further emphasizing the importance of comprehensive PRH approaches.

Alongside the impact in QoL, emotional distress is the most common psychological symptom in cancer patients, irrespective of disease stage, primary cancer site and phase of treatment (39). Brain tumor patients however experience even higher rates of emotional distress and mental health conditions compared to those with non-CNS tumors (11,12). Emotional distress is not only a QoL key determinant but also a factor that can influence clinical outcomes. For example, prospective cohort studies indicate that cancer patients experiencing the highest levels of distress have higher mortality rates compared to those with the lowest distress levels, across multiple cancer types (10). Furthermore, Rucińska & Osowiecka (40), highlighted that PRH programs contribute to both physical and psychological well-being, leading to reduced complications, shorter hospital stays, and improved mental health outcomes. Consistent with these findings, our study demonstrated that participants in the Prehabilita project experienced a significant reduction in emotional distress, suggesting that PRH can be an effective intervention for addressing the psychological challenges faced by brain tumor patients. Despite previous studies not specifically focusing on neuro-oncology patients, they emphasize the role of PRH in mitigating emotional distress across cancer populations. Our study results add to these findings by incorporating both quantitative psychological assessments and qualitative questionnaires, thus providing a more comprehensive understanding of the impact of PRH on EWB in brain tumor patients.

From a clinical perspective, baseline data showed that over half of participants (13 out of 25) scored above the HADS clinical threshold of 15, according with Spanish validation studies (41). While the mean baseline score was just below the cutoff, individual variability indicated significant emotional distress in a substantial portion of the sample. After the intervention, the number of participants above the clinical threshold dropped to 10, suggesting that the improvement was not only statistically significant but also clinically meaningful. Still, some gains remained within the subclinical range, reflecting partial yet relevant improvements. Additionally, the observed sex-by-time interaction, showing a significant reduction in distress among women, should be interpreted with caution. Given the reduced number of studies on PRH in the neuro-oncology population, it remains unclear whether there are sex differences in its overall effectiveness. Nonetheless, psychological evidence suggests that men and women may respond differently depending on the specific type of psychological intervention. According to (42), women tend to derive greater benefit from supportive therapies characterized by empathy, affiliation, and emotional expressiveness, while men may respond more effectively to interpretive approaches. This evidence implies that the women-driven effects on EWB observed during PRH may be attributed to differences in how individuals engage with supportive interventions, rather than reflecting a disparity in overall treatment efficacy. Such an interpretation aligns with the supportive nature of PRH, which is particularly consistent with the type of intervention to which women show higher responsiveness.

### 4.2 Psychological mechanisms behind EWB improvement: the potential role of control attitudes

Results for the secondary measures show an increase in the perception of control over the disease among participants after completing PRH. This suggests that individuals may feel more empowered and in control of their own health outcomes following the intervention. These results are consistent with findings from Powell and colleagues (15), who noted that cancer patients reported a heightened sense of control over their health and treatment outcomes following PRH. Similarly, other studies on non-brain cancers have also highlighted the role of control attitudes in shaping psychological outcomes. Research by Sharif (17) and Hjörleifsdóttir et al. (18) found that patients with higher levels of perceived control, or an internal locus of control, exhibited lower levels of depression and anxiety, along with an improved QoL at the time of their cancer diagnosis. Our results support the notion that PRH programs can also promote psychological empowerment in brain cancers, through the enhancement of information, support, and social factors, which are essential elements of patient-centered care. While these findings emphasize the potential benefits of fostering a sense of control, it remains unclear how this translates to brain tumor patients, given the distinct cognitive and neurological challenges they may face. In our study, while we found a clear increase in control attitudes after PRH, there was no direct association between it and changes in QoL or emotional distress. This could be explained by our limited sample size, compared to previous similar work. However, it might also indicate that the mechanisms through which PRH enhances well-being may be more complex and multifaceted, potentially involving various psychological constructs beyond mere perception of control. In this vein, the discrepancy between our present study findings, and those of previous research by Sharif (17) and Hjörleifsdóttir et al. (18) may indicate that PRH in brain cancer patients enhances the perception of control, but this perception alone does not necessarily translate into reductions in anxiety or depression or improved QoL.

Remarkably, our results also show no changes in perceived stress after PRH. This is consistent with previous findings by Scriney and colleagues (43), who reviewed the effectiveness of PRH in improving psychological and functional outcomes in cancer patients aged 18–55 years and found that psychological PRH interventions did not significantly alter stress. This observation raises important implications regarding the multifaceted nature of psychological interventions in the context of cancer treatment. While PRH appears to enhance certain psychological attributes, such as the sense of control over one’s medical circumstances, it does not necessarily lead to a direct decrease in stress levels. Importantly, the mere presence of stressors, such as cancer diagnosis or the rigors of treatment, constructs the environmental context in which resilience is both required and developed. Stressors are integral to resilience because they provide the challenges that individuals must navigate and adapt to, thereby fostering growth and psychological fortitude. This perspective aligns with Arenaza-Urquijo & Vemuri (44), who describe resilient individuals as those who engage in adaptive coping strategies rather than eliminating stress altogether. Similarly, Gillis et al. (45) highlight that PRH interventions—incorporating exercise, nutrition, and psychosocial support—enhance physiological reserve and functional capacity, ultimately fostering resilience. Their findings suggest that patients who undergo PRH exhibit greater resilience, leading to better recovery outcomes.

### 4.3 Qualitative results

The qualitative findings of this study highlight several key themes related to the EWB of patients participating in a PRH program, with particular attention to the support received and the perceived control over their medical situation. Two main analytical themes emerged from the data: (1) Sources of emotional support during PRH; (2) control over medical situation.

We found that access to information and a better understanding of their medical situation played a crucial role in empowering patients during PRH. These findings are further aligned with the ones that Powell et al. (15) found in their qualitative study. Indeed, in this previous study, many participants highlighted that having detailed information and the opportunity to ask questions significantly reduced their emotional distress. Our quantitative results further support this notion, demonstrating that informed patients experience greater EWB. In fact, knowing what to expect not only alleviated emotional distress but also contributed to their EWB by providing a sense of “security” and “control.”

Another important aspect that extends beyond perceived control is the influence of broader support systems, including social support from family, peers, PRH staff, and healthcare professionals, as well as coping strategies such as emotional reassurance and the feeling of being accompanied. Our findings are aligned with those of (46) who indicated that promoting supportive networks is critical for resilience, as it helps mitigate the adverse effects of stress and trauma by facilitating emotional support and connection. In our study, some participants emphasized the importance of feeling “accompanied” and “safe,” underscoring how strong social support networks positively influence their EWB. Notably, PRH support staff may not only provide essential medical guidance but also serve as key figures within this support system, offering reassurance and continuity throughout the process. Altogether, our results highlight the significance of both emotional and informational support in enhancing the EWB of patients undergoing PRH. The findings further suggest that a multifaceted support system plays a crucial role in helping patients navigate the emotional challenges of the PRH process.

Therefore, given the established relationship between emotional distress and lower QoL and poorer clinical outcomes in patients with brain tumors (8,10), interventions that enhance social support and promote coping strategies that foster a sense of control might offer substantial benefits. Reducing emotional distress improves treatment tolerance, adherence to medical recommendations, and overall QoL (47). Thus, our findings suggest that perceived control is one key factor, alongside social support and coping strategies that provide emotional reassuranceIdentifying these determinants of EWB can help refine PRH strategies, potentially enhancing not only surgical outcomes but also broader clinical results, including patient survival. Strengthening social support networks and fostering adaptive coping skills may be valuable targets for future clinical interventions.

## 5. Conclusions

Non-invasive PRH in neuro-oncology appears to enhance EWB prior to surgery, as indicated by improvements in QoL and emotional distress measures. These effects appear more pronounced in women. Moreover, control attitudes emerged as a potential contributor to EWB improvement, possibly enhancing resilience throughout the surgical process. However, qualitative analyses suggest that additional factors, such as social network support and access to information, may also play a critical role. Altogether, these findings underscore the importance of investigating and integrating psychological variables and targeted interventions into PRH programs, particularly in neuro-oncology, where EWB is significantly impacted.

### Limitations

This investigation is not without limitations. The absence of a control group makes it difficult to confirm the effects of the treatment from natural progression patterns. Additionally, the small sample size, particularly for secondary measures during the telephonic enrichment phase, and the study’s design not focusing on these outcomes, resulted in limited data on EWB and related psychological factors. Therefore, larger studies with more comprehensive psychological assessments are needed to confirm the generalizability of these findings in PRH programs for neuro-oncology patients.

## Data availability

To protect participant confidentiality and in accordance with ethical approval and informed consent, the datasets from this study are not publicly accessible. Access is restricted and may be granted upon request. For further details, please contact the corresponding author.

## Ethics approval and consent to participate

All procedures from the present study were performed in accordance with the Helsinki declaration. The study was approved by the Research Ethical Committee of Fundació Unió Catalana d’Hospitals (approval number: CEI 21/65, version 1, 13/07/2021). Clinician participants provided informed consent by reviewing and agreeing to a consent statement presented by the research team, prior to responding to any questions. Verbal informed consent was obtained from patient participants prior to the interview. Interviews were not audio-recorded but were transcribed in real time.

## Consent for publication

Participants in the study provided consent for the use of anonymized quotations in research publications. All identifying details have been excluded to ensure participant confidentiality.

## Author’s contributions

N.B.B. contributed to investigation, formal analysis, data curation, writing – original draft and editing, and visualization. J.M.T., K.A.P., and A.P.L. contributed to conceptualization and methodology, investigation, writing – review and editing, and resources. S.D.G. and A.R.V. contributed to investigation, data curation, and writing – review and editing. K.A.P and A.R.V contributed to project administration and supervision. E.B.O., R.P.A., C.H.A., and D.B.F. contributed to investigation and writing – review and editing. All authors read and approved the final manuscript.

## Acknowledgements

**Prehabilita Working Group:** Núria Bargalló; Leonardo Boccuni; Roger Bragulat; Mònica Buxeda-Rodríguez; María Cabello-Toscano; Gerardo Conesa-Bertrán; César Garrido; Mireia Illueca-Moreno; Carlos Laredo; David Leno-Colorado; Ruth Lau; Estela Lladó-Carbó; Jesús Martín-Fernández; Francisco Martínez-Ricarte; Emma Muñoz-Moreno; Cristóbal Perla Y Perla; José Carlos Pariente; Maria Redondo-Camós; and Gloria Villalba-Martínez.

## Declaration of generative AI and AI-assisted technologies in the writing process

During the preparation of this work the authors used [OpenAI. *GPT-4.5* model] to support the organization and clustering of codes of the qualitative analysis with shared meanings into broader themes.

After using this tool/service, the authors reviewed and edited the content as needed and take full responsibility for the content of the publication.

## List of abbreviations

CAS: Control Attitudes Scale
CNS: Central Nervous System
DLPFC: Dorsolateral Prefrontal Cortex
EORTC – QLQ: European Organization for Research and Treatment of Cancer Quality of Life Questionnaire
EWB: Emotional Well-Being
HADS: Hospital Anxiety Depression Scale
MRI: Magnetic Resonance Imaging
NIBS: Non-Invasive Brain Stimulation
NICP: Neuromodulation-Induced Cortical Prehabilitation
PRH: Prehabilitation
PSS-4: 4-item Perceived Stress Scale
QoL: Quality of life
TMS: Transcranial Magnetic Stimulation

## References

1. Fan Y, Zhang X, Gao C, Jiang S, Wu H, Liu Z, et al. Burden and trends of brain and central nervous system cancer from 1990 to 2019 at the global, regional, and country levels. Arch Public Health. 2022 Sept 17;80(1):209.

2. Bray F, Laversanne M, Sung H, Ferlay J, Siegel RL, Soerjomataram I, et al. Global cancer statistics 2022: GLOBOCAN estimates of incidence and mortality worldwide for 36 cancers in 185 countries. CA Cancer J Clin. 2024 May;74(3):229–63.

3. Red Española de Cáncer. Estimaciones de la incidencia del cancer en España, 2024. 2024 [cited 2025 July 28]. REDECAN. Available from: https://redecan.org/en

4. Angom RS, Nakka NMR, Bhattacharya S. Advances in Glioblastoma Therapy: An Update on Current Approaches. Brain Sci. 2023 Oct 31;13(11):1536.

5. Chanbour H, Chotai S. Review of Intraoperative Adjuncts for Maximal Safe Resection of Gliomas and Its Impact on Outcomes. Cancers. 2022 Jan;14(22):5705.

6. Gerritsen JKW, Zwarthoed RH, Kilgallon JL, Nawabi NL, Versyck G, Jessurun CAC, et al. Impact of maximal extent of resection on postoperative deficits, patient functioning, and survival within clinically important glioblastoma subgroups. Neuro-Oncol. 2023 May 1;25(5):958–72.

7. Pertz M, Schlegel U, Thoma P. Sociocognitive Functioning and Psychosocial Burden in Patients with Brain Tumors. Cancers. 2022 Jan;14(3):767.

8. Randazzo D, Peters KB. Psychosocial distress and its effects on the health-related quality of life of primary brain tumor patients. CNS Oncol. 2016 Oct 31;5(4):241–9.

9. Jimenez-Labaig P, Aymerich C, Braña I, Rullan A, Cacicedo J, González-Torres MÁ, et al. A comprehensive examination of mental health in patients with head and neck cancer: systematic review and meta-analysis. JNCI Cancer Spectr. 2024 Apr 30;8(3):pkae031.

10. Grimmett C, Heneka N, Chambers S. Psychological Interventions Prior to Cancer Surgery: a Review of Reviews. Curr Anesthesiol Rep. 2022 Jan 31;12(1):78–87.

11. Otto-Meyer S, Lumibao J, Kim E, Ladomersky E, Zhai L, Lauing KL, et al. The interplay among psychological distress, the immune system, and brain tumor patient outcomes. Curr Opin Behav Sci. 2019 Aug;28:44–50.

12. Liu F, Huang J, Zhang L, Fan F, Chen J, Xia K, et al. Screening for distress in patients with primary brain tumor using distress thermometer: a systematic review and meta-analysis. BMC Cancer. 2018 Dec;18(1):124.

13. Ji W, Sang C, Zhang X, Zhu K, Bo L. Personality, Preoperative Anxiety, and Postoperative Outcomes: A Review. Int J Environ Res Public Health. 2022 Sept 26;19(19):12162.

14. Fehrenbach MK, Brock H, Mehnert-Theuerkauf A, Meixensberger J. Psychological Distress in Intracranial Neoplasia: A Comparison of Patients With Benign and Malignant Brain Tumours. Front Psychol. 2021 Aug 16;12:664235.

15. Powell R, Davies A, Rowlinson-Groves K, French DP, Moore J, Merchant Z. Impact of a prehabilitation and recovery programme on emotional well-being in individuals undergoing cancer surgery: a multi-perspective qualitative study. BMC Cancer. 2023 Dec 14;23(1):1232.

16. Brahmbhatt P, Sabiston CM, Lopez C, Chang E, Goodman J, Jones J, et al. Feasibility of Prehabilitation Prior to Breast Cancer Surgery: A Mixed-Methods Study. Front Oncol. 2020 Sept 25;10:571091.

17. Pahlevan Sharif S. Locus of control, quality of life, anxiety, and depression among Malaysian breast cancer patients: The mediating role of uncertainty. Eur J Oncol Nurs. 2017 Apr;27:28–35.

18. Elisabet Hjörleifsdóttir, Eva Charlotte Halapi, Þórhalla Sigurðardóttir. Health Locus of Control and Psychological and Somatization Disorder in Icelandic Outpatients with Cancer: A Quantitative Study | Nordicum-Mediterraneum [Internet]. 2024 [cited 2025 July 28]. Available from: https://nome.unak.is/wordpress/volume-19-no-1-2024/new-article-double-blind-peer-review-volume-19-no-1-2024/health-locus-of-control-and-psychological-and-somatization-disorder-in-icelandic-outpatients-with-cancer-a-quantitative-study/

19. Boccuni L, Roca-Ventura A, Buloz-Osorio E, Leno-Colorado D, Delgado-Gallén S, Cabello-Toscano M, et al. Non-invasive prehabilitation to foster widespread fMRI cortical reorganization before brain tumor surgery: lessons from a case series. J Neurooncol [Internet]. 2024 July 23 [cited 2024 Aug 22]; Available from: 10.1007/s11060-024-04774-4

20. Boccuni L, Abellaneda-Pérez K, Martín-Fernández J, Leno-Colorado D, Roca-Ventura A, Prats Bisbe A, et al. Neuromodulation-induced prehabilitation to leverage neuroplasticity before brain tumor surgery: a single-cohort feasibility trial protocol. Front Neurol. 2023 Oct 2;14:1243857.

21. Boccuni L. Exploring the neural basis of non-invasive prehabilitation in brain tumour patients: An fMRI-based case report of language network plasticity.

22. Aaronson NK, Ahmedzai S, Bergman B, Bullinger M, Cull A, Duez NJ, et al. The European Organization for Research and Treatment of Cancer QLQ-C30: a quality-of-life instrument for use in international clinical trials in oncology. J Natl Cancer Inst. 1993 Mar 3;85(5):365–76.

23. Chow R, Lao N, Popovic M, Chow E, Cella D, Beaumont J, et al. Comparison of the EORTC QLQ-BN20 and the FACT-Br quality of life questionnaires for patients with primary brain cancers: a literature review. Support Care Cancer. 2014 Sept;22(9):2593–8.

24. Zigmond AS, Snaith RP. The Hospital Anxiety and Depression Scale. Acta Psychiatr Scand. 1983 June;67(6):361–70.

25. Moser DK, Dracup K. Psychosocial recovery from a cardiac event: The influence of perceived control. Heart Lung. 1995 July;24(4):273–80.

26. Moser DK, Riegel B, McKinley S, Doering LV, Meischke H, Heo S, et al. The Control Attitudes Scale-Revised: Psychometric Evaluation in Three Groups of Patients With Cardiac Illness. Nurs Res. 2009 Jan;58(1):42–51.

27. Cohen S, Kamarck T, Mermelstein R. A Global Measure of Perceived Stress. J Health Soc Behav. 1983 Dec;24(4):385.

28. Vallejo MA, Vallejo-Slocker L, Fernández-Abascal EG, Mañanes G. Determining Factors for Stress Perception Assessed with the Perceived Stress Scale (PSS-4) in Spanish and Other European Samples. Front Psychol. 2018 Jan 26;9:37.

29. Krishnasamy M, Hassan H, Jewell C, Moravski I, Lewin T. Perspectives on Emotional Care: A Qualitative Study with Cancer Patients, Carers, and Health Professionals. Healthcare. 2023 Feb 4;11(4):452.

30. OpenAI. GPT-4.5 model [Internet]. 2025. Available from: https://openai.com

31. Lefaucheur JP, Aleman A, Baeken C, Benninger DH, Brunelin J, Di Lazzaro V, et al. Evidence-based guidelines on the therapeutic use of repetitive transcranial magnetic stimulation (rTMS): An update (2014–2018). Clin Neurophysiol. 2020 Feb;131(2):474–528.

32. Cash RFH, Zalesky A, Thomson RH, Tian Y, Cocchi L, Fitzgerald PB. Subgenual Functional Connectivity Predicts Antidepressant Treatment Response to Transcranial Magnetic Stimulation: Independent Validation and Evaluation of Personalization. Biol Psychiatry. 2019 July 15;86(2):e5–7.

33. Okamoto M, Dan H, Sakamoto K, Takeo K, Shimizu K, Kohno S, et al. Three-dimensional probabilistic anatomical cranio-cerebral correlation via the international 10–20 system oriented for transcranial functional brain mapping. NeuroImage. 2004 Jan 1;21(1):99–111.

34. Fitzgerald PB, Maller JJ, Hoy KE, Thomson R, Daskalakis ZJ. Exploring the optimal site for the localization of dorsolateral prefrontal cortex in brain stimulation experiments. Brain Stimulat. 2009 Oct;2(4):234–7.

35. Cash RFH, Weigand A, Zalesky A, Siddiqi SH, Downar J, Fitzgerald PB, et al. Using Brain Imaging to Improve Spatial Targeting of Transcranial Magnetic Stimulation for Depression. Biol Psychiatry. 2021 Nov;90(10):689–700.

36. Lobo DN, Pavel Skořepa, Gomez D, Greenhaff PL. Prehabilitation: high-quality evidence is still required. Br J Anaesth. 2023 Jan;130(1):9–14.

37. Yun-Jen Chow, Shiow-Ching Shun. Cancer Prehabilitation Programs and Their Effects on Quality of Life. Oncol Nurs Forum [Internet]. 2018 Nov 1 [cited 2025 July 28]; Available from: https://onf.ons.org/onf/45/6/cancer-prehabilitation-programs-and-their-effects-quality-life

38. McIsaac DI, Kidd G, Gillis C, Branje K, Al-Bayati M, Baxi A, et al. Relative efficacy of prehabilitation interventions and their components: systematic review with network and component network meta-analyses of randomised controlled trials. BMJ. 2025 Jan 22;e081164.

39. Grassi L, Caruso R, Riba MB, Lloyd-Williams M, Kissane D, Rodin G, et al. Anxiety and depression in adult cancer patients: ESMO Clinical Practice Guideline. ESMO Open. 2023 Apr;8(2):101155.

40. Rucińska M, Osowiecka K. Prehabilitation as an extra approach to usual care for cancer patients. Nowotw J Oncol. 2022 Oct 19;72(5):294–302.

41. Herrero MJ, Blanch J, Peri JM, De Pablo J, Pintor L, Bulbena A. A validation study of the hospital anxiety and depression scale (HADS) in a Spanish population. Gen Hosp Psychiatry. 2003 July;25(4):277–83.

42. Ogrodniczuk JS, Piper WE, Joyce AS, McCallum M. Effect of patient gender on outcome in two forms of short-term individual psychotherapy. J Psychother Pract Res. 2001;10(2):69–78.

43. Scriney A, Russell A, Loughney L, Gallagher P, Boran L. The impact of prehabilitation interventions on affective and functional outcomes for young to midlife adult cancer patients: A systematic review. Psychooncology. 2022 Dec;31(12):2050–62.

44. Arenaza-Urquijo EM, Vemuri P. Resistance vs resilience to Alzheimer disease: Clarifying terminology for preclinical studies. Neurology. 2018 Apr 10;90(15):695–703.

45. Gillis C, Ljungqvist O, Carli F. Prehabilitation, enhanced recovery after surgery, or both? A narrative review. Br J Anaesth. 2022 Mar;128(3):434–48.

46. Sippel LM, Pietrzak RH, Charney DS, Mayes LC, Southwick SM. How does social support enhance resilience in the trauma-exposed individual? Ecol Soc. 2015;20(4):art10.

47. Samman RR, Timraz JH, Mosalem Al-Nakhli A, Haidar S, Muhammad Q, Irfan Thalib H, et al. The Impact of Brain Tumors on Emotional and Behavioral Functioning. Cureus [Internet]. 2024 Dec 8 [cited 2025 July 28]; Available from: https://www.cureus.com/articles/313929-the-impact-of-brain-tumors-on-emotional-and-behavioral-functioning

