## Supplementary Material for "The impact of non-invasive prehabilitation before surgery on emotional well-being in neuro-oncology patients: Insights from the Prehabilita project"

### Materials and Methods

#### Supplementary file 1. Full interview guide

1. Did you receive support to cope with stress or to help you if your mood was affected?
2. How did you find this support? (For example, how did you feel about it? How useful did you find it?)
3. Do you feel that you have a greater sense of control over your medical situation after attending Prehabilita?

#### Supplementary file 2. Audit trail (qualitative thematic analysis)

#### 1. Overview of dataset

| Item | Description |
| --- | --- |
| Number of interviews | 9 |
| Participant profile | Patients who underwent prehabilitation (PRH program) |
| Data collection period | December 2024 – February 2025 |
| Data format | Verbatim transcripts from structured interviews |

#### 2. Coding process

| Step | Description |
| --- | --- |
| Initial read-through | Conducted to note the overall tone, emotional content, and unexpected patterns. |
| Line-by-line coding | Codes were generated inductively from the text using manual annotation, supported by AI tools (OpenAI, 2025). |
| Number of initial codes | 12 |
| Coding memo example | “Several participants mention receiving strong guidance from medical professionals (medical doctors, nurse practitioners, psychologists, rehabilitation teams).” |

**3. Development of themes**

| Code Group | Emerging Theme | Notes |
|---|---|---|
| “Spouses, children, close relatives providing emotional strength.” | Family support | |
| “Receiving guidance from doctors, nurses, psychologists, rehabilitation teams.” | Perceived support from professionals | |
| “Feeling supported, accompanied, serenity, safe, comfort.” | Emotional and psychological support | Overlap with emotional well-being themes |
| “Receiving help from colleagues, friends, and social circles.” | Community and social support | |
| “Adaptation, calmness, strength, and fear” | Psychological adaptation | Grouped based on the emotional process of coping |
| “Information explained vaguely” | Need for more clear information | Captures concerns about unclear communication from professionals |
| “Reduced sensation of ‘no control’; information, prevention, and serenity indicate that understanding their condition helped them gain confidence” | Confidence and understanding | Theme reflects the link between being informed and feeling emotionally prepared |

**4. Decision log**

| Date | Decision | Justification |
|---|---|---|
| Feb 20, 2025 | Split general theme “Support” into four distinct sub-themes: Family Support, Healthcare Team Support, Community and Social Support, and Emotional and Psychological Support | Participants clearly distinguished between different sources and types of support. Splitting allowed for more nuanced analysis and representation. |
| Feb 21, 2025 | Dropped “Need for More Clear Information” as a separate standalone theme, though remarks were noted in context | This concern was mentioned by only one participant and lacked enough depth for theme status. Incorporated as a subtheme in broader thematic interpretation. |
| Feb 24, 2025 | Merged support-related sub-themes into a broader overarching category: | Although expressed differently, these sub-themes shared a common emotional function and |

| | | |
|---|---|---|
| | Sources of Emotional Support | were consolidated for clarity in final reporting. |
| Feb 24, 2025 | Refined theme “Psychological Adaptation” to include coping-related descriptors such as adaptation, calmness, strength, and fear | These emotional descriptors consistently emerged in responses as part of patients’ efforts to manage uncertainty and stress, especially during preparation for surgery. |
| Feb 26, 2025 | Created new theme “Confidence and Understanding” from codes such as information, control, serenity, and prevention | These concepts consistently clustered around the idea that greater understanding of their medical condition gave participants confidence and a sense of control. |

**Supplementary file 3. Control analysis - Euclidean distances**

To examine the potential influence of stimulation site location on treatment response, we calculated the Euclidean distance between each participant’s TMS stimulation coordinate and three widely used left dorsolateral prefrontal cortex (DLPFC) reference points: (1) the group-derived site from Cash et al. (35) [(x = –41, y = 43, z = 27)], (2) the anatomical location proposed by Okamoto et al. (33) [(x = –43, y = 58, z = 40)], and (3) the coordinate used by Fitzgerald et al. (34) [(x = –46, y = 45, z = 38)]. Pearson’s product-moment correlations were performed to assess whether distance from each reference site was associated with changes in EWB, as measured by the HADS, EORTC-QLQ, and EORTC-BN20 scales.

No significant associations were found between distance to any of the three DLPFC reference sites and changes in emotional well-being (EWB), as assessed by Spearman’s rho. Correlations between Euclidean distance and HADS score changes were $\rho = -0.22$, $p = .281$ for Cash et al. (32), $\rho = -0.22$, $p = .281$ for Okamoto et al. (33), and $\rho = -0.22$, $p = .281$ for Fitzgerald et al. (34). Across all sites and measures (HADS, EORTC-QLQ, and EORTC-BN20), no associations reached statistical significance. Thirteen out of 25 participants (52%) showed EWB improvement regardless of stimulation site distance.

| DLPFC Reference Coordinate | EWB Measure | Spearman's ρ (rho) | *p*-value | Effect Size (Fisher's *z*) | SE (z) |
|---|---|---|---|---|---|
| **Cash et al. (32)** | HADS | –0.22 | .28 | –0.22 | 0.21 |
| | EORTC-QLQ | –0.03 | .89 | 0.03 | 0.22 |
| | EORTC-BN20 | 0.00 | 1.00 | 0.00 | .022 |
| **Okamoto et al. (33)** | HADS | –0.23 | .28 | –0.23 | 0.21 |
| | EORTC-QLQ | –0.09 | .69 | –0.09 | 0.22 |
| | EORTC-BN20 | 0.04 | 0.86 | 0.04 | .022 |
| **Fitzgerald et al. (34)** | HADS | –0.27 | .19 | –0.28 | 0.21 |
| | EORTC-QLQ | 0.11 | .60 | 0.12 | 0.22 |
| | EORTC-BN20 | 0.05 | 0.81 | 0.05 | .022 |

**Table S1.** Correlation Between Distance to DLPFC Reference Coordinates and Changes in Emotional Well-Being (EWB). Note: Euclidean distance was computed between each participant's TMS stimulation site and each of the three DLPFC coordinates. Change scores reflect differences from TP1 (pre-prehabilitation) to TP2 (post-prehabilitation). Emotional well-being (EWB) was assessed using HADS, EORTC-QLQ, and EORTC-BN20. None of the correlations reached statistical significance.
